# Number of interventional clinical trials registered per million population per country worldwide

**DOI:** 10.1101/2024.11.22.24317736

**Authors:** Jonas Leth Bjerg, Dimitrinka Nikolova, Christian Gluud

**Affiliations:** Copenhagen Trial Unit, Centre for Clinical Intervention Research, The Capital Region, Copenhagen University Hospital ⍰ Rigshospitalet, Copenhagen, Denmark; Cochrane Hepato-Biliary Group, Copenhagen Trial Unit, Centre for Clinical Intervention Research, The Capital Region, Copenhagen University Hospital ⍰ Rigshospitalet, Copenhagen, Denmark; Department of Regional Health Research, The Faculty of Health Sciences, University of Southern Denmark, Odense, Denmark

## Abstract

**Background:** Historically, Sweden and Denmark have excelled in clinical trial publications per million population, but many trials have remained unpublished. Enforceable policies for trial registration before launch, made global and national clinical research assessments possible. The World Health Organisation (WHO) hosts the International Clinical Trials Registry Platform (ICTRP) for interventional clinical trials, with data from 1999.

**Objective:** To find out if the retrospective or prospective trial registration activities per million population per country during the 2009 to 2022 period have improved after establishing the number of interventional clinical trials registered per million population per country during the 1999 to 2022 period.

**Methods:** We used the WHO ICTRP to extract the numbers of registered interventional clinical trials. We entered data in Microsoft Excel to analyse two separate periods, 1999 to 2022 and 2009 to 2022. The second period aimed at filtering out the pickup of the registration requirements. To obtain the trial registration activity by country, we divided the number of interventional clinical trials registered by the country’s population in millions. Information on population numbers originated from the World Bank, the Central Intelligence Agency World Factbook, and The National Institute of Statistics and Economic Studies (of France) as of 2022. We collected data as of 03-01-2024.

**Results:** In February 2023, there were 943.113 interventional clinical trials on the ICTRP. After excluding 32.309 trials with unknown country of origin, 910.804 trials from 229 countries and territories were analysed. For countries or territories with populations over 1 million, the top five in interventional clinical trial registrations per million during 1999-2022 were Denmark, Estonia, Belgium, The Netherlands, and Latvia. For 2009-2022, they were Denmark, Estonia, Belgium, The Netherlands, and New Zealand. The top five overall, irrespective of population, for both periods were Pitcairn, The Vatican, Tokelau, Niue, and Denmark.

**Conclusions:** Since 1999, there has been worldwide growth in interventional trial registrations, especially among top-contributing countries. The global average stands at approximately 115 registered trials per million people from 1999 to 2022, highlighting the need for continued efforts to improve trial registration and production. Smaller Western countries lead in interventional clinical trials per million population.

## Background

The assessment of countries’ contribution to global clinical research was previously conducted by evaluating the number of articles published on clinical trials [1, 2]. From 1945 until 2005, the USA and UK were in the forefront of publications on randomised clinical trials in total [1], while Sweden and Denmark were in front with numbers as high as 891 and 864 publications on randomised clinical trials per million population during that same period of time [1]. In 2011 the topic on number of publications was revisited, and results were confirmatory [2]. The results are of interest, but due to the observation that about every second randomised clinical trial is never published [3], a national research activity may not become properly gauged. Hence, we found it important to also assess the number of interventional clinical trials registered per country during more recent years.

The negative impact of publication bias has led to global worries about how this type of clinical research waste could be stopped [4-7]. The Food and Drug Administration Modernization Act section 113 (FDAMA 113) was introduced in USA in 1997 [8] demanding protocol registration before recruitment of participants in trials concerning serious or life-threatening diseases [9]. It was one of the early steps towards proper trial registration aiming to secure a bias-free publication. In 2004 the International Committee of Medical Journal Editors (ICMJE) issued a statement acknowledging the selective reporting of trials, resulting in the ICMJE demanding trial registration at or before the onset of enrolment of participants by July 2005 [10]. In 2005, at the Ministerial Summit on Health Research Mexico, the World Health Organization (WHO) called upon the global scientific community, international partners, the private sector, civil society, and other relevant stakeholders to *“establish a platform linking clinical trials registers in order to ensure a single point of access and the unambiguous identification of trials”* [11]. These actions together with activities like the AllTrials intitiative, The Cochrane Collaboration, and many others, have been of great importance to oppose publication bias and promote an increase in registrations of unfinished or abandoned trials due to mostly unsatisfactory results [12-16].

The present study assesses all registered interventional clinical trials in the WHO International Clinical Trial Registry Platform (ICTRP), enabling the possibility to bring forward the individual country’s trial registration per million population.

### Aims

Our primary aim was to assess the number of interventional clinical trials registered per million population per country during the 1999 to 2022 period. Our secondary aim was to find out if the retrospective or prospective trial registration activities per million population per country during the 2009 to 2022 period have improved.

## Methods

### Data sources and data extraction

The ICTRP, established in 2005, gathers world-wide data on clinical trials registrations from the WHO primary and partner registries (22 in total), data providers, and registries working with the ICTRP towards becoming primary registries (https://www.who.int/clinical-trials-registry-platform/network). The ICTRP database contains numbers on the yearly registered interventional clinical trials for the period 1999 to 2022[17]. We extracted the number of interventional clinical trial registrations by country by selecting the ‘interventional clinical trials’ term from a list of options, followed by searches for 1) trial registrations during the years 1999 to 2022 (the whole available period), and 2) only for the years 2009 to 2022. The year 2009 was selected allowing the countries time to adopt to the ICMJE request for registration from 2005 [10]. We selected the time periods by marking the wanted period in chart A on the website, and then obtaining the numbers on countries from chart E [17]. We copied the numbers from the database into an Excel worksheet, enabling the possibility to obtain a visual representation when comparing the numbers of registered interventional clinical trials with populations in different countries. We used Excel to divide the number of interventional clinical trials by the respective country’s population in millions to obtain the number of registered trials per million population. JB, one of the authors, contacted WHO by email on 25-03-2024 to request spreadsheets from the ICTRP, containing further relevant information for our study, but such were not received. Therefore, the extracted data are obtained solely from the WHO ICTRP.

We gathered data on population sizes for each country, primarily from The World Bank [18] and The Central Intelligence Agency (CIA) World Factbook [19]. We found data for the French colonies at The National Institute of Statistics and Economic Studies (of France) (French Guiana, Guadeloupe, Martinique, Mayotte, Réunion) [20].

### Data analysis

We planned two rankings based on country population. The first included only countries with more than 1 million population. The second included all countries and territories. It must be noted that due to the smallness of some countries and territories, their metric of number of registered interventional clinical trials per year per million population is multiplied by 1 million, and therefore, the numbers may be inflated and less precise.

We handled all numbers and graphs in Microsoft Excel.

## Results

### Description of search and selection

There were 943.113 registered interventional clinical trials on the WHO ICTRP in February 2023. After excluding 32.309 trials with unknown country of origin, we were left with a total of 910.804 trials, with trial registrations belonging to 229 countries and territories. JB entered the data for analysis in Excel, in conformity with our methods section. DN performed a random data check to validate the correctness of data entry. JB calculated the registration activity of each country by dividing the amount of interventional clinical trials registered with the country’s population and multiplying by 1.000.000.

Dividing the total number of interventional clinical trial registrations (n = 910.804) by the world population in millions (n = 7.9 billion in 2022), gave a global mean of 115 trials for the 1999 to 2022 period and 96.7 trials for the 2009 to 2022 period (n = 766.115).

### Top countries with populations of 1 million population or above

Figure 1 and Figure 2 show the top 20 countries with the highest number of registrations of interventional clinical trials among countries with a population of 1 million and above for the two periods in separate. During the 1999 to 2022 period, Denmark, Estonia, Belgium, The Netherlands, and Latvia were the leading countries. During the 2009 to 2022 period, Denmark, Estonia, Belgium, The Netherlands, and New Zealand were the leading countries.

**Figure 1.**
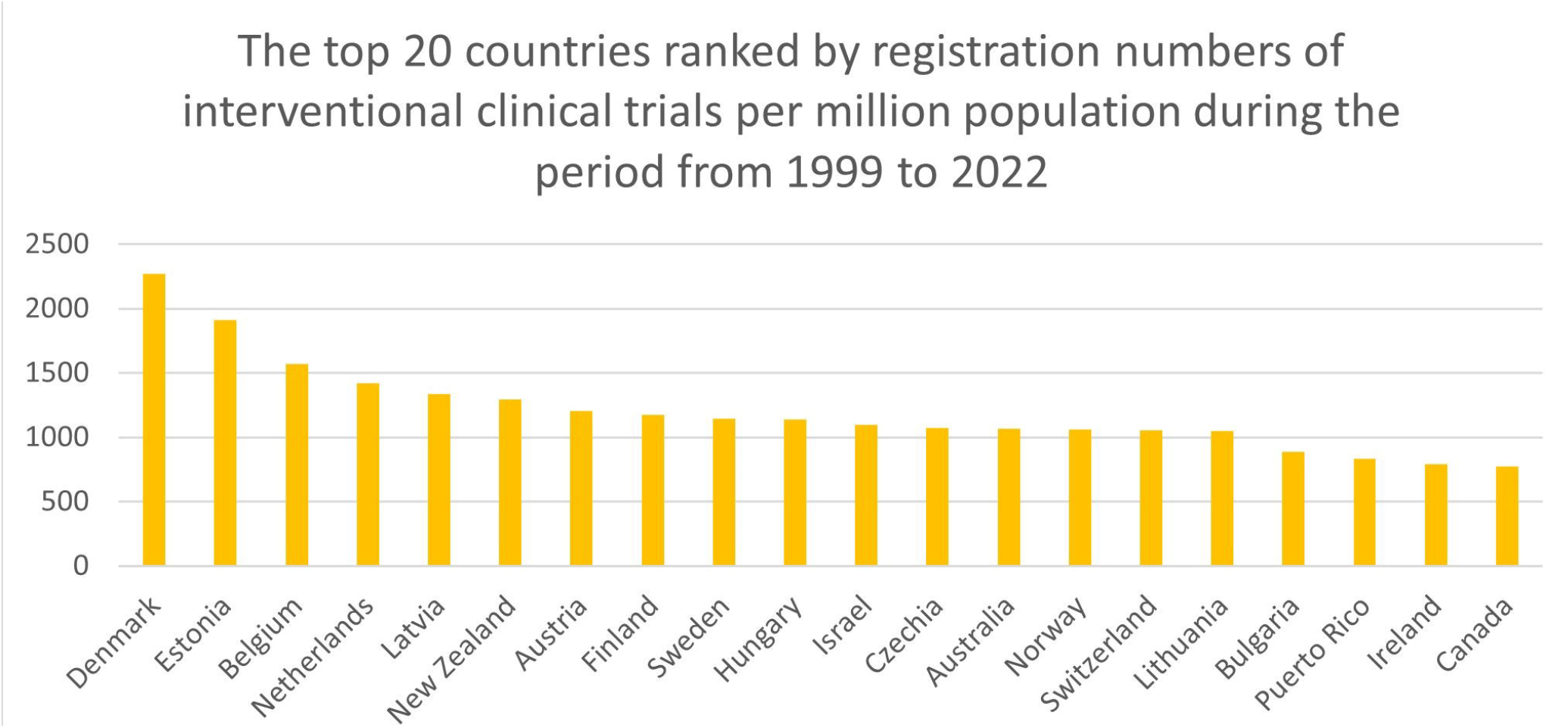
The top 20 countries ranked by registration numbers of interventional clinical trials per million population during the period from 1999 to 2022

**Figure 2.**
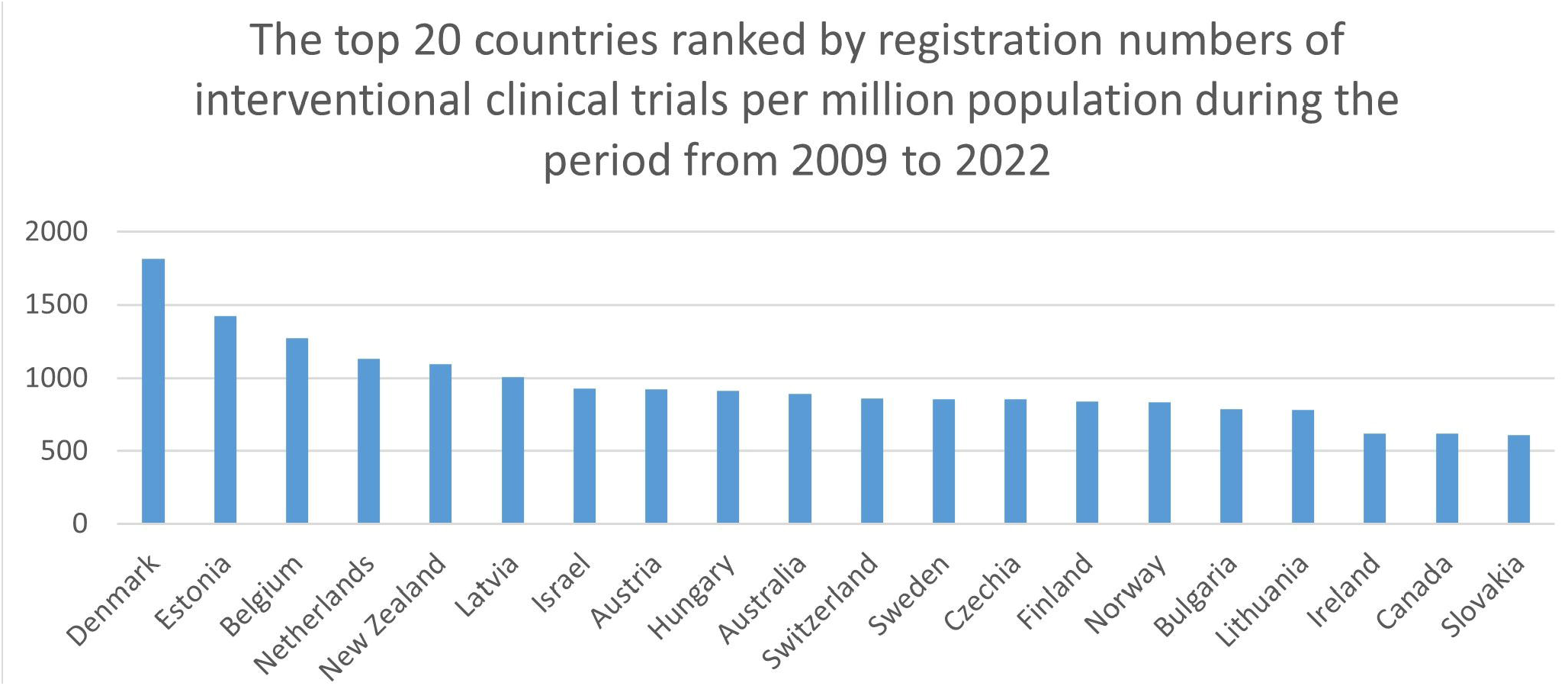
The top 20 countries ranked by registration numbers of interventional clinical trials per million population during the period from 2009 to 2022

The number of registered interventional clinical trials in Denmark during the 1999 to 2022 period was 2267.8 per million population and during the 2009 to 2022 period it was 1815.7 per million population. For Estonia, the number of registered interventional clinical trials during the 1999 to 2022 period was 1910.5 per million population and during the 2009 to 2022 period it was 1420.5 per million population. And for Belgium, the number of registered interventional clinical trials during the 1999 to 2022 period was 1568.4 per million population and during the 2009 to 2022 it was 1270.6 per million population.

Supplementary table 1 shows the number of interventional clinical trials per million population in the top 20 countries with a population of 1 million or more.

### Top countries among all countries and territories

We also produced an overview of the top 20 countries and territories during the same two periods, 1999 to 2022 and 2009 to 2022. See Figure 3 and Figure 4. Leading countries in both time periods were Pitcairn (territory of UK), The Vatican, Tokelau (territory of New Zealand), Niue (self-governing in free association with New Zealand), and Denmark. Supplementary table 2 shows the number of registered interventional clinical trials per million population in the top 20 countries and territories, irrespective of population size.

**Figure 3.**
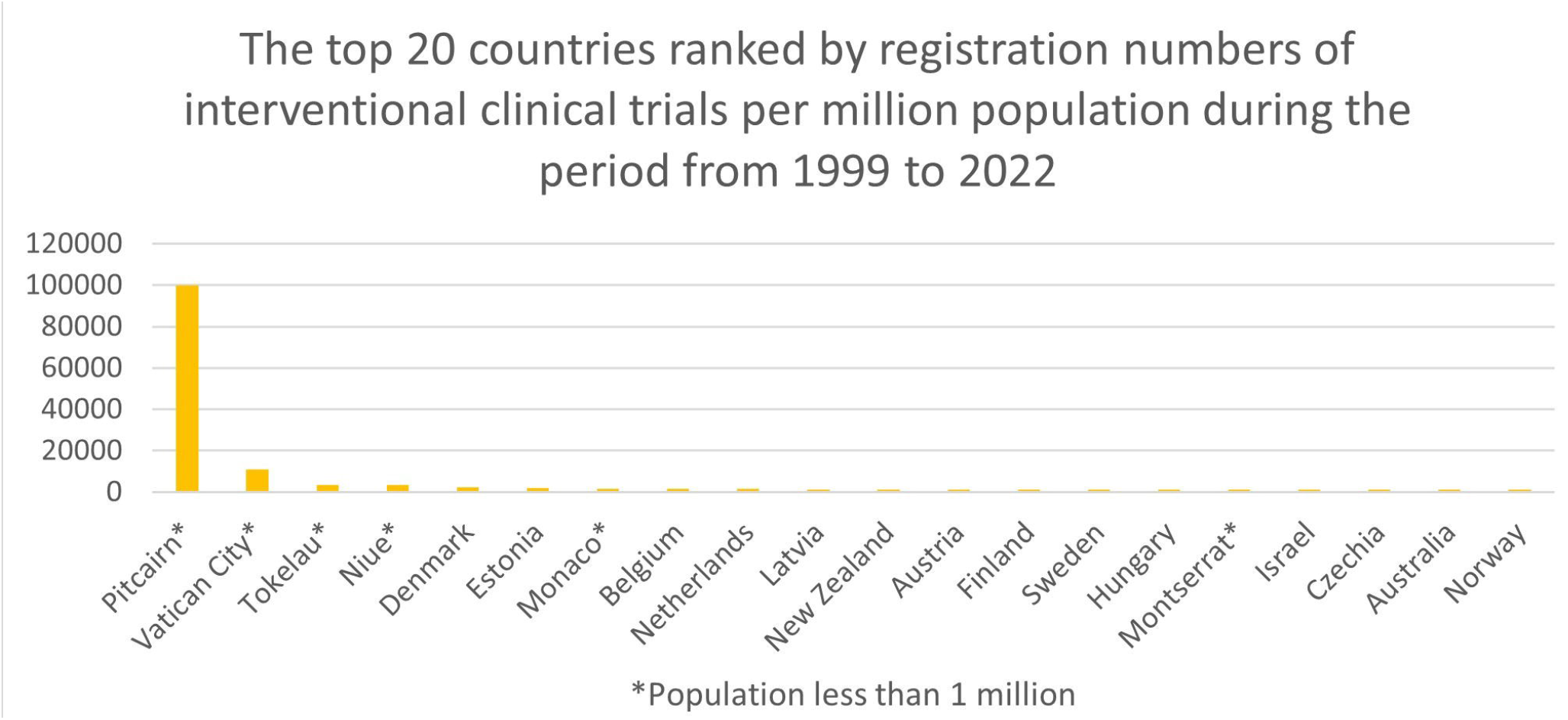
The top 20 countries ranked by registration numbers of interventional clinical trials per million population during the period from 1999 to 2022 ^*^Population less than 1 million

**Figure 4.**
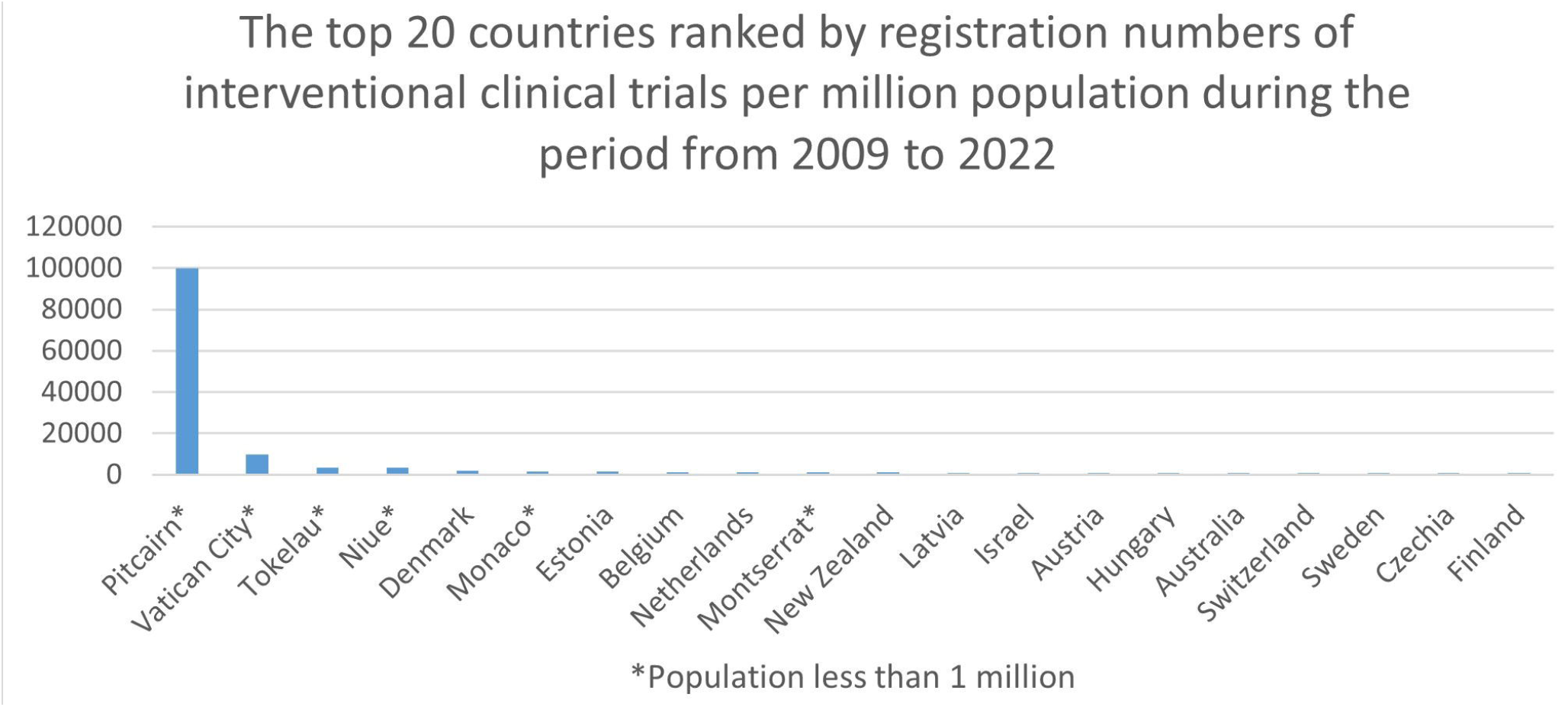
The top 20 countries ranked by registration numbers of interventional clinical trials per million population during the period from 2009 to 2022 ^*^Population less than 1 million

For the period of 1999 to 2022, the results we obtained on the number of registered interventional clinical trials were as follows: for Pitcairn, 100.000 trials per million population (5 trials and 50 people); for the Vatican, 11.000 trials per million population (11 trials and 1000 people); and for Tokelau, 3643 trials per million population (6 trials and 1647 people).

For the period 2009 to 2022, the results we obtained on the number of registered interventional clinical trials were as follows: for Pitcairn, no change; for the Vatican, it was 10.000 per million population; and for Tokelau, no change.

Data on all countries, present in the ICTRP, is available in supplementary table 3.

## Discussion

### Summary of findings

When not accounting for small population countries (< 1 million), Denmark came out as the country with highest number of trial registrations, according to our unit of measurement. For all countries and territories, Pitcairn was without competition the top contributor of interventional clinical trials when the number of registered trials was held against its population (50 people in 2021). In both scenarios, with and without countries with smaller populations, it becomes clear that the ranking of forefront countries is unchanged in both time intervals.

### Limitations and strengths

This study aimed to paint a picture of the registration of interventional clinical trials by incorporating numbers on all countries, but with no analysis of each trial’s scope, design, and utility. This results in a limitation where our results do *not* rely on factors, such as sample size and trial quality. To consider the scale and quality of each trial would be important because a high number of trial registrations per million population, not taking in consideration sample size and trial quality, could result in increased production of smaller scale trials [2].

Second, limitations in this study do also repeat the limitations of the ICTRP. Some registered trials had missing data, resulting in a relatively larger number of trials with unknown country of origin (n = 32.309). This corresponded to less than 5% of the interventional clinical trials, and we considered the missing data spread evenly across all countries, not affecting the overall result. As we looked at the number of registered interventional clinical trials divided by a country’s population in millions, we encountered a misrepresentation when a population was too small. Therefore, we used a lower limit of 1 million population, highlighting results of 69 countries (6 of which are present in Figure 3 and Figure 4).

Third, our data on country populations are a snapshot of a certain point in time and are not representative for the entire 1999 to 2022 period. Though the world population is growing [21], some countries, especially in eastern Europe, are experiencing population decline [22].

Therefore, one should be aware that some of the mentioned forefront countries could credit their number of registered interventional trials per million population to a population decrease as this would result in an increased position. Similarly, countries who have had an increase in population would obtain a lower number of registered interventional trials per million population.

This study followed our aims and present results based on the net registration of interventional clinical trials. Sufficiently powered trials are with a higher value than insufficiently powered trials, and one could ponder whether countries with large populations, like China or USA, have registered fewer but larger trials compared to small top countries such as Denmark. This is worth investigating further.

As mentioned, this study aimed to include all available data on trial registration from every country in the world, also counting colonies or territories with independent data (n = 229). It is hard to determine whether we have succeeded in this, but we recognise the strength in using the data made available by the WHO through their ICTRP.

### Results in context

The results presented in this paper show that there have been countries consistently in the forefront of trial production, but also that previously unacknowledged countries have risen, becoming some of the top contributors.

Countries like Estonia, Czechia, Lithuania, Bulgaria, Puerto Rico, and Ireland have gone from not even making the top 20 list in the period 1946-2005 [1], to setting an example that others should aim for today. Especially Estonia has made a big progress during the last 23 years. While formerly being at the very top of contributors, Sweden has fallen a few places in the top 20 ranking compared with the results in the two articles based on publication numbers of randomised and controlled trials [1, 2]. Though not to neglect that Sweden, among others, is still one of the big contributors on the international scene of interventional clinical trials.

Though Denmark has been recognised as the best performing country in our article in terms of clinical trial registrations, it has still much to improve. Based on a report published in TranspariMED [23] in February 2024, the Scandinavian countries had lack of publishing trial results. Denmark, at the time of the report, still had 19% unpublished trial results for trials registered in the period 2016 to 2019. Even though countries have a high registration and production of interventional clinical trials, the number could be higher. Countries like Denmark should naturally aim for a 100% publication of trial results, as this activity is most importantly following ethical principles for medical research involving humans (https://www.wma.net/policies-post/wma-declaration-of-helsinki-ethical-principles-for-medical-research-involving-human-subjects/).

Highly populated areas are China with 1.412.175.000 population, India with 1.417.173.170 population, and the continent of Africa with 1.332.006.610 population. These populations together correspond to about 50% of our global population. Looking at the interventional clinical trial registrations per million population during the 1999 tot 2022 period (China = 45; India = 29; and Africa = 17), it is obvious that there is an imbalance in the distribution of trial registrations as the world mean is 115 registered interventional clinical trials per million population.

### Conclusions

Looking at our two timeframes of measurement, we expected to see a big increase in registered trials produced in the period 2009 to 2022, and so we did. The three forefront countries Denmark, Estonia, and Belgium, all had a big increase in trial production in the mentioned period, with Denmark registering 10.718 interventional clinical trials in the 2009 to 2022 period from a total of 13.387 in the 1999 to 2022 period. The same tendency was seen with Estonia (n = 1.916 in 2009 to 2022, n = 2.577 in 1999 to 2022) and Belgium (n = 14.848 in 2009 to 2022, n = 18.328 in 1999 to 2022). This increase in registration of trials could be a result of the common advancement and development in health research, but we may also conclude that this increase proceeds from the ICMJE’s requirement of trial registration along with other initiatives. Though unregistered trials are still present [24], we are nevertheless seeing an increase in registrations. Large countries, and the largest economies, like USA, China, and Japan [25], had the overall highest production of interventional clinical trials, but these numbers tend to fade into smaller proportions after undergoing our method of analysis. This emphasises the importance of a standardised approach and incentives for trial data sharing.

Since 1999, the number of interventional clinical trial registrations have been increasing globally, and for many countries which are among the top contributors, the registrations have increased almost exponentially. During the twenty-three-year period that our study examines, several countries have managed to register thousands of interventional clinical trials. Whatever the reason for this increase in registrations is, e.g. ICMJE recommendations, it is an increase of major importance for obtaining more complete data on healthcare interventions [26]. In 2007, a benchmark of 700 trials per million population per country was suggested, which would have resulted in millions of trials 60 years onwards [1]. Currently, almost 20 years after the suggested benchmark, the number of registered interventional clinical trials per million worldwide is 115 (i.e. the total number of registered trials (n = 910.804) divided by the world population in millions). This shows that even though we are seeing an increase in trial registration by country, we must keep pursuing methods for a more thorough trial registration and production.

## Data Availability

All data produced in the present study are available upon reasonable request to the authors

## Declarations

### Ethics approval and consent to participate

not applicable

### Consent for publication

not applicable

### Availability of data and materials

all data generated or analysed during this study are included in this published article [and its supplementary information files].

## Authors’ contributions

JB acted as lead author. DM carried out a data check, and DM and CG helped with structure and content of the study. All authors critically reviewed the manuscript for important content and approved the final version of the manuscript. The guarantor of this study is JB.

## Acknowledgements

not applicable

